# From Symptom to Outcome: Defining Clinically Meaningful Patient-Reported Appetite Loss in Non-Small-Cell Lung Cancer

**DOI:** 10.1101/2025.07.09.25331191

**Authors:** Jiawei Zhou, Benyam Muluneh, Quefeng Li, Lynne I. Wagner, Yuchen Wang, Jim H. Hughes

**Author notes:** **Corresponding Author:** Jiawei Zhou, University of North Carolina at Chapel Hill, 2320 Kerr Hall, Chapel Hill, NC, 27599, USA.

## Abstract

**Background:** Appetite loss is a common and distressing symptom in non-small-cell lung cancer (NSCLC), driven by both treatment side effects and disease progression. It often leads to unintended weight loss and cancer cachexia, significantly impairing patients’ quality of life and survival. Yet, appetite loss remains under-recognized in oncology care, with no standard assessment tools or universal management guidelines. In this study, we developed a predictive model to identify clinically meaningful thresholds of appetite loss that were associated with significant weight reduction and decreased survival. We aim to highlight the clinical consequences of appetite loss and advocate for more patient-centered treatment strategies that address this often overlooked but impactful symptom in cancer care.

**Methods:** We analyzed longitudinal patient-reported appetite scores, measured by the Lung Cancer Symptom Scale (LCSS), and body weight data from 476 NSCLC patients receiving supportive care and recovering from prior chemoradiotherapy (recovering cohort). A mechanism-based population modeling approach was used to predict the impact of appetite changes on body weight, accounting for significant data variability and missingness. Model validation was conducted using data from 380 NSCLC patients undergoing docetaxel chemotherapy (chemotherapy cohort). Clinically meaningful appetite loss thresholds were determined based on the model-predicted appetite loss associated with a 3.5 kg body weight loss and significantly worse survival (p < 0.05).

**Results:** We found that, according to the LCSS (100 mm visual analogue scale), a 30 mm improvement in appetite corresponded to a 3.5 Kg weight gain in the recovering cohort, while a 23 mm decline correlated with a 3.5 Kg weight loss in the chemotherapy cohort. Significant associations were observed between appetite loss trajectories and overall survival. Clinically meaningful thresholds for appetite loss were identified as 4 mm at 1 month and 11 mm at 3 months, both significantly associated with reduced OS in patients receiving docetaxel chemotherapy.

**Conclusions:** We developed a population predictive model to characterize the relationship between patient-reported appetite and body weight, identifying clinically meaningful thresholds for appetite loss. This work highlights the importance of managing appetite loss in oncology care and supports its use as a quantitative endpoint in clinical trials and practice.

## Introduction

Lung cancer remains the leading cause of cancer-related mortality in the United States, accounting for an estimated 124,730 deaths in 2025. [1] Non-small-cell lung cancer (NSCLC) accounting for more than 80% of all lung cancer cases and the five-year survival rate is only 26.4%. [2] In addition to poor prognosis, NSCLC patients often experience a high symptom burden—including cough, pain, fatigue, and appetite loss—driven by both the disease and treatment-related toxicity. These symptoms significantly impair their quality of life and daily functioning.[3]

Among these symptoms, appetite loss is particularly prevalent and distressing, affecting approximately 60%–70% of lung cancer patients. [4] Persistent appetite loss can lead to unintentional weight loss, malnutrition, and cancer cachexia–a syndrome characterized by severe muscle wasting, metabolic abnormalities, and increased mortality. [5, 6] Despite its high prevalence and clinical consequences, appetite loss remains under-recognized and under-managed in routine oncology practice. There are currently no standardized assessment tools or universally accepted evidence-based guidelines for managing appetite loss. [5] Patients often report that this symptom significantly impacts their well-being and quality of life, yet it receives relatively little attention in clinical care. [7, 8]

This gap underscores an urgent need to better understand how appetite loss relates to downstream clinical outcomes, including weight reduction and overall survival, and to develop targeted strategies to mitigate its impact. In this study, we address this unmet need by evaluating patient-reported appetite loss during treatment in NSCLC and quantifying its relationship with body weight change and survival outcomes.

In our study, we focus on patient-reported appetite loss rather than clinician-reported adverse events, as it more accurately reflects patient experience and aligns with the growing emphasis on patient-centered care in oncology. [9–11] However, patient-reported outcomes (PRO) data present unique analytical challenges, including high variability and frequent missingness. [12] To address these challenges, we proposed a novel population modeling approach. Population modeling is a computational method used to find common patterns in data from many people, while also accounting for between-subject variability and measurement noise. It is widely used in pharmaceutical research to understand how medicines behave in different patients. [12] In our prior work, we demonstrated that this approach could handle the variability and noise in the PRO data and predict survival based on patient-reported symptoms. [13]

In the current study, we extended this approach to model the mechanistic relationship between patient-reported appetite and body weight using data from NSCLC patients. We further explored how changes in appetite are associated with patient overall survival (OS). We aim to identify clinically meaningful thresholds of appetite loss associated with significant weight loss and worse survival outcomes. These thresholds may serve as early indicators of cancer cachexia risk. By linking subjective symptoms with objective outcomes, our study provides a quantitative framework to guide appetite monitoring and symptom management in NSCLC treatment. Ultimately, our goal is to highlight the clinical consequences of appetite loss and advocate for more patient-centered treatment strategies that address this often overlooked but impactful symptom in cancer care.

## Methods

### Ethic Statement

This retrospective analysis used fully de-identified data. The study was granted an Institutional Review Board (IRB) exemption by the University of North Carolina at Chapel Hill.

### Data Source and Study Description

#### Recovering study cohort

Appetite–body weight model was developed using data from a Phase III randomized, double-blind clinical trial (NCT00409188). [14] This trial enrolled patients with unresectable stage III NSCLC who had completed chemoradiotherapy 4–12 weeks prior to randomization and had achieved either stable disease or an objective response. Participants included in our analysis were from the control arm, receiving supportive care such as psychosocial support and nutritional support, and were in the recovery phase following chemoradiation.

#### Chemotherapy study cohort

To validate the model and workflow, we used data from another Phase III clinical trial (NCT00532155). [15] This trial assessed the efficacy of aflibercept combined with docetaxel versus docetaxel alone in patients with advanced or metastatic nonsquamous NSCLC. Participants included in our analysis were from the control arm and received docetaxel at 75 mg/m², administered intravenously over one hour every three weeks, along with best supportive care.

Both trials were conducted in accordance with the International Conference on Harmonisation Good Clinical Practice (ICH-GCP) guidelines and the Declaration of Helsinki. Study protocols were approved by local regulatory authorities and institutional ethics committees, and all participants provided written informed consent prior to enrollment.

### Data Exclusions and Imputations

All control arm participants from both cohorts were included in the analysis. For each participant, patient-reported Lung Cancer Symptom Scale (LCSS) scores [16], longitudinal body weight data, baseline demographics, clinical characteristics, and overall survival outcomes were extracted. The patient-reported LCSS consists of nine items measured using visual analogue scales (0–100 millimeters [mm]) to assess lung cancer symptom burden and quality of life. [17, 18] In our analysis, we focused on the appetite-related item, “*How is your appetite?*”, where a score of 0 mm indicates “*As good as it could be*” and 100 mm indicates “*As bad as it could be.*” Higher scores represent more severe appetite loss.

In the recovering study cohort, LCSS questionnaires were collected at baseline, weeks 2, 5, and 8, then every 6 weeks from week 13 until disease progression, with follow-up assessments at 6 and 12 weeks post-progression and every 12 weeks thereafter. Body weight was measured at baseline, weekly during weeks 1–8, and every 6 weeks from week 13 until progression. [14] In the chemotherapy study cohort, LCSS questionnaires were collected at baseline, cycle 2, cycle 4, and at the end of treatment. Body weight was measured at every treatment cycle. [15]

Participants with only one appetite or body weight measurement were excluded from data analysis. No imputations were performed for missing data.

### Appetite**–**Body Weight Model Development

To characterize the relationship between patient-reported appetite and body weight, we developed a mechanism-based model using longitudinal data from the recovering study cohort. The model captures underlying pharmacological mechanisms and disease progression in appetite loss and body weight reduction and consists of two components: the trajectory of appetite scores and their impact on body weight.

In the appetite model, patients in the recovery cohort showed early improvement in appetite as they recovered from chemoradiotherapy-related side effects. As this early improvement could not be separated from any placebo effects [19], both recovery and placebo were described using an asymptotic exponential with *PMAX* and *Kp* parameters. The appetite loss due to disease progression was modeled using a separate exponential function, governed by a rate parameter, *SLP*. A higher score indicates greater appetite loss; therefore, appetite improvement components were modeled as reductions from the baseline, while appetite loss components were added to the baseline. The mathematical model for the appetite trajectory is presented in **Equation 1**.

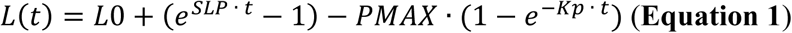

Where *L(t)* is the appetite scores change over time, *L0* is the appetite score at baseline, *SLP* is appetite loss rate, *PMAX* is the maximal appetite improvements, and *Kp* is the appetite improvements offsite rate (1/day).

Changes in appetite were linked to changes in body weight, with the assumptions that appetite loss inhibits body weight gain through decreased food intake. An indirect response model was utilized to account for the physiological lag between changes in appetite and the corresponding changes in body weight. [20] The mathematical model for the appetite–body weight relationship is presented in **Equation 2**.

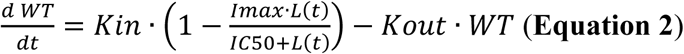

Where *WT* is body weight over time, *L(t)* is the appetite scores change over time (same as **Equation 1**), *Kin* is the constant rate of body weight increase, *Kout* is the first-order rate constant of body weight reduction, *Imax* is the maximum appetite loss impact on body weight reduction, *IC50* is the appetite loss that causes 50% *Imax*.

Additionally, *Kout* parameter was set as **Equation 3** to ensure a constant body weight if appetite doesn’t change over time.

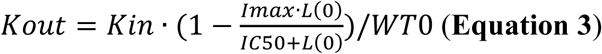

Where *WT0* is the body weight at baseline and *L(0)* is the appetite score at baseline.

### Population Modeling Approach

Given the substantial variability in the patient-reported appetite scores, we used a population modeling approach to estimate the model parameters. This approach allows us to estimate parameter values that represent the average behavior of the overall population, in addition to values for each individual that capture how each participant’s response differs from the average within the study cohort. [12, 21] We also tested whether covariates (demographics and baseline clinical characteristics) could help explain differences in appetite and body weight patterns between subjects. [22] To check model performance, we used goodness-of-fit plots and visual predictive checks (VPCs). [23, 24] More details about this population modeling approach are provided in the **Supplementary Methods**.

### Model Validation

We further validated the appetite–body weight model using data from the chemotherapy cohort. The model structure and parameters describing the relationship between appetite and body weight (*Kin*, *Imax*, and *IC50* in **Equation 2**) remained the same as the recovery cohort. However, parameters for the appetite trajectory (**Equation 1**) and baseline body weight (*WT0*) were re-estimated. These decisions assumed that while the effects of appetite on body weight are expected to be consistent across studies, the trajectory of appetite and body weight may not be the same, due to different study designs and participant populations.

### Model Simulation

To evaluate the appetite–body weight relationship in NSCLC population, the final model was used to simulate 1,000 virtual studies, varying the model estimates according to parameter precision. Using the simulated appetite–body weight trajectories, we estimated the degree of appetite loss required to achieve a 3.5 kilogram (Kg) weight reduction—equivalent to 5% of body weight for a 70 Kg individual, a commonly used threshold in cancer cachexia trials [25], based on the chemotherapy cohort model parameters. Similarly, we estimated the amount of appetite improvements associated with a 3.5 Kg weight gain using the recovering cohort model parameters. Separate simulations were performed using individual parameters for each subject in the dataset to capture patient-specific appetite and body weight trajectories. These results were used to assess the associations between appetite loss and survival outcomes.

### Associations Between Appetite Trajectory and Survival

To assess the relationship between appetite trajectory and OS, we developed Cox proportional hazards models that incorporated both prognostic clinical characteristics and key appetite–body weight model parameters (*L0*, *PMAX*, *SLP*, *WT0*, and *Imax*). The relative contribution of each parameter to survival was visualized using forest plots. Prognostic clinical variables were identified using the Least Absolute Shrinkage and Selection Operator (LASSO) algorithm, with five-fold cross-validation. Variables were selected based on the λ value within one standard error of the minimum to ensure model robustness.

### Clinically Meaningful Appetite Loss

Clinically meaningful appetite loss was defined as the level of appetite loss significantly associated with OS. Using model simulations from the chemotherapy study cohort, we obtained appetite scores change from baseline for each individual patient at 1 and 3 months after chemotherapy. Patients were then grouped by their appetite loss scores using cutoffs ranging from 0 mm to 15 mm. Cox proportional hazards models were constructed to compare the two groups, adjusting for confounding clinical variables selected by LASSO. The lowest cutoff yielding p-value < 0.05 was identified as the clinically meaningful threshold. A parallel analysis was conducted using body weight loss cutoffs to determine the level of weight reduction significantly associated with OS.

### Statistical Analysis

Appetite trajectory parameter comparisons between the recovering and chemotherapy cohorts were performed using Wilcoxon tests. Differences in OS between groups were evaluated using Kaplan–Meier curves and compared with log-rank tests. Associations between appetite trajectory parameters or appetite cutoffs with OS were evaluated using Cox proportional hazards models, with hazard ratio (HR), 95% confidence intervals (CI), and p-value reported. A two-sided p-value < 0.05 was considered statistically significant.

### Software and Code Availability

Population appetite–body weight model was developed using Monolix 2024R1. The model simulations were performed using Simulx 2024R1. Both Monolix and Simulx could be downloaded at https://lixoft.com/products/. Plots and survival analyses were performed using R 4.4.1 and RStudio Version 2022.07.1+554. The figures were reorganized in GraphPad Prism (Version 10.4.1). The model codes are provided in the **Supplementary Codes**.

### Data Availability

The data were accessed through Project Data Sphere under license in accordance with the Project Data Sphere Data Use Agreement. [26] The raw analysis data are available from the corresponding author upon reasonable request.

## Results

### Study Data and Workflow

Data from two study cohorts (recovering and chemotherapy) were incorporated in our analysis and the study workflow is illustrated in **Figure 1A**. The recovering cohort included 476 participants, with 6,517 patient-reported appetite scores and 4,284 body weight measurements. The chemotherapy cohort included 380 participants, contributing 1,179 appetite scores and 3,698 body weight observations. (**Figure S1**) Participant demographics, baseline clinical characteristics, and survival data are summarized in **Table 1**. Longitudinal trajectories of individual appetite scores and body weight changes from baseline are shown in **Figure S2A-D**. In the recovering cohort, a substantial proportion of participants experienced weight gain (**Figure S2B**), whereas most patients in the chemotherapy cohort showed weight loss over time (**Figure S2D**).

**Figure 1.**
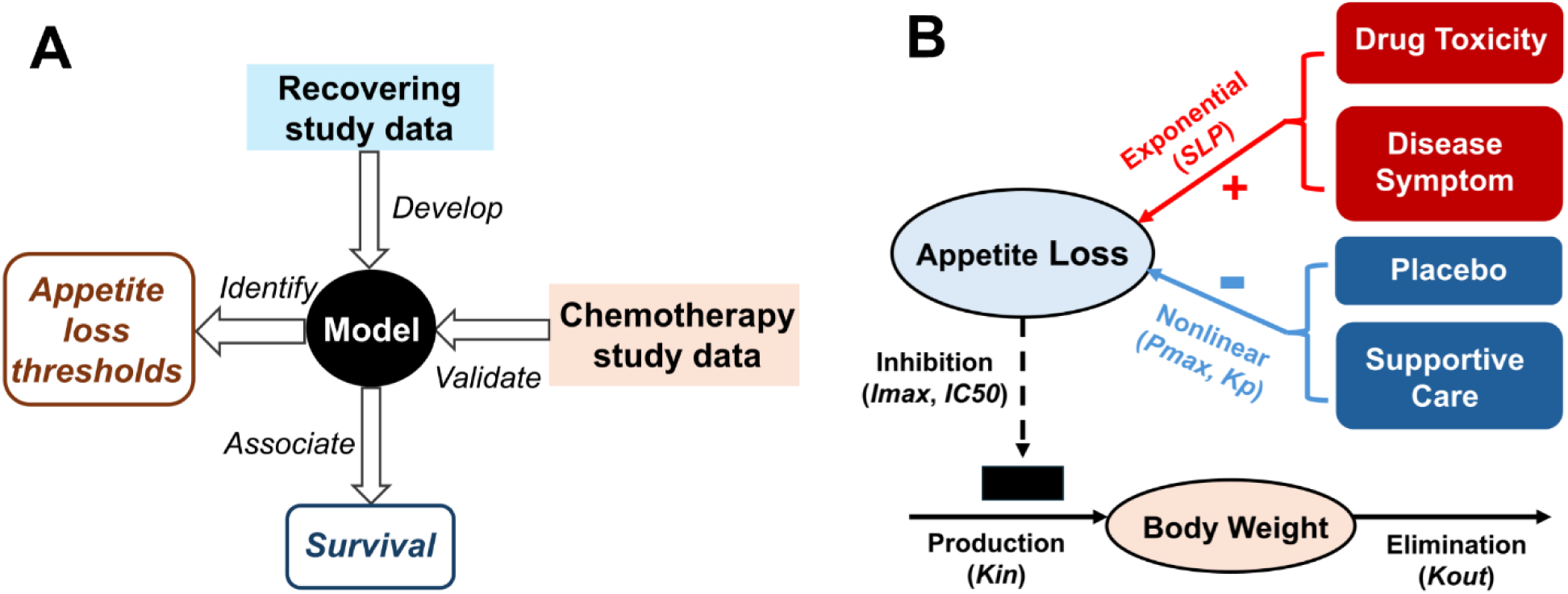
Study workflow and appetite–body weight model diagram. (A) The appetite–body weight model was developed using recovering study data and validated using chemotherapy study data. The model was used to identify clinically meaningful appetite loss thresholds and the associations between appetite loss and survival. (B) Appetite loss during treatment is influenced by both drug-related toxicity and disease-related symptoms, while placebo effects and supportive care can help improve appetite. In turn, reduced appetite limits body weight gain.

**Table 1.** Demographics of two study populations.

| **Trial** | **Recovering (N = 476)** | **Chemotherapy (N = 380)** |
| --- | --- | --- |
| **Intervention** | Placebo and supportive care | Docetaxel |
| **Age, years** | 61 (24, 83) | 61 (27, 80) |
| **Sex** |  |  |
| Male | 324 (68.1%) | 243 (63.9%) |
| Female | 152 (31.9%) | 137 (36.1%) |
| **Race** |  |  |
| White/Caucasian | 441 (92.6%) | 333 (87.6%) |
| Other | 35 (7.4%) | 47 (12.4%) |
| **Height, cm** | 171 (144, 194) | 168 (140, 193) |
| Missing | 67 (14.1%) | 0 (0%) |
| **Weight, Kg** | 74 (38.6, 140.8) | 71 (35, 118.7) |
| Missing | 69 (14.5%) | 3 (0.8%) |
| **Anxiety/Depression history** |  |  |
| Yes | 39 (8.2%) | 39 (10.3%) |
| No | 437 (91.8%) | 341 (89.7%) |
| **Smoking status** |  |  |
| Yes | 443 (93.1%) | 80 (21.1%) |
| No | 33 (6.9%) | 94 (24.7%) |
| Former smoker | Not Applicable | 206 (54.2%) |
| **ECOG performance status** |  |  |
| 0 | 201 (42.2%) | 130 (34.2%) |
| 1 | 275 (57.8%) | 232 (61.1%) |
| 2 | 0 (0%) | 18 (4.7%) |
| **Overall Survival, months** | 21 (1.7, 65.7) | 11.7 (1.6, 36.1) |
| Missing | 92 (19.3%) | 9 (2.4%) |
| **Disease stage** |  |  |
| I | 0 (0%) | 24 (6.3%) |
| II | 0 (0%) | 10 (2.6%) |
| III | 476 (100%) | 111 (29.2%) |
| IV | 0 (0%) | 223 (58.7%) |
| Missing | 0 (0%) | 12 (3.2%) |
| **Response to initial**<br>**chemoradiotherapy** |  | Not Applicable |
| Objective response | 332 (69.7%) |  |
| Stable disease | 144 (30.3%) |  |
| **Type of initial**<br>**chemoradiotherapy** |  | Not Applicable |
| Concurrent | 289 (60.7%) |  |
| Sequential | 183 (38%) |  |
| Missing | 6 (1.3%) |  |

Data are number (%) or median (range), unless otherwise specified.
ECOG, Eastern Cooperative Oncology Group.

### Characterizing Appetite**–**Body Weight Relationship in NSCLC

The appetite–body weight model structure is illustrated in **Figure 1B**, where appetite loss due to drug toxicity and disease symptoms was modeled using an exponential function, while a separate asymptotic exponential component captured supportive care and placebo-related improvements. Appetite loss, in turn, inhibited body weight gain via an indirect response model. Population modeling approach was used to estimate typical model parameters for the overall population, between-subject variability, covariate effects, and residual error based on data from the recovering cohort. Final model parameters are listed in **Table S1**. The appetite–body weight model adequately described the data, as shown by the alignment between observed and predicted values for appetite scores and body weight in **Figure 2A–B**. VPC of body weight change over time showed that observations data were well captured by model simulations. (**Figure 2C**) **Figure 2D** presents model predictions overlaid with observations in six randomly selected subjects, demonstrating good alignment. These diagnostics confirm that the model appropriately captured the data from the recovering cohort.

**Figure 2.**
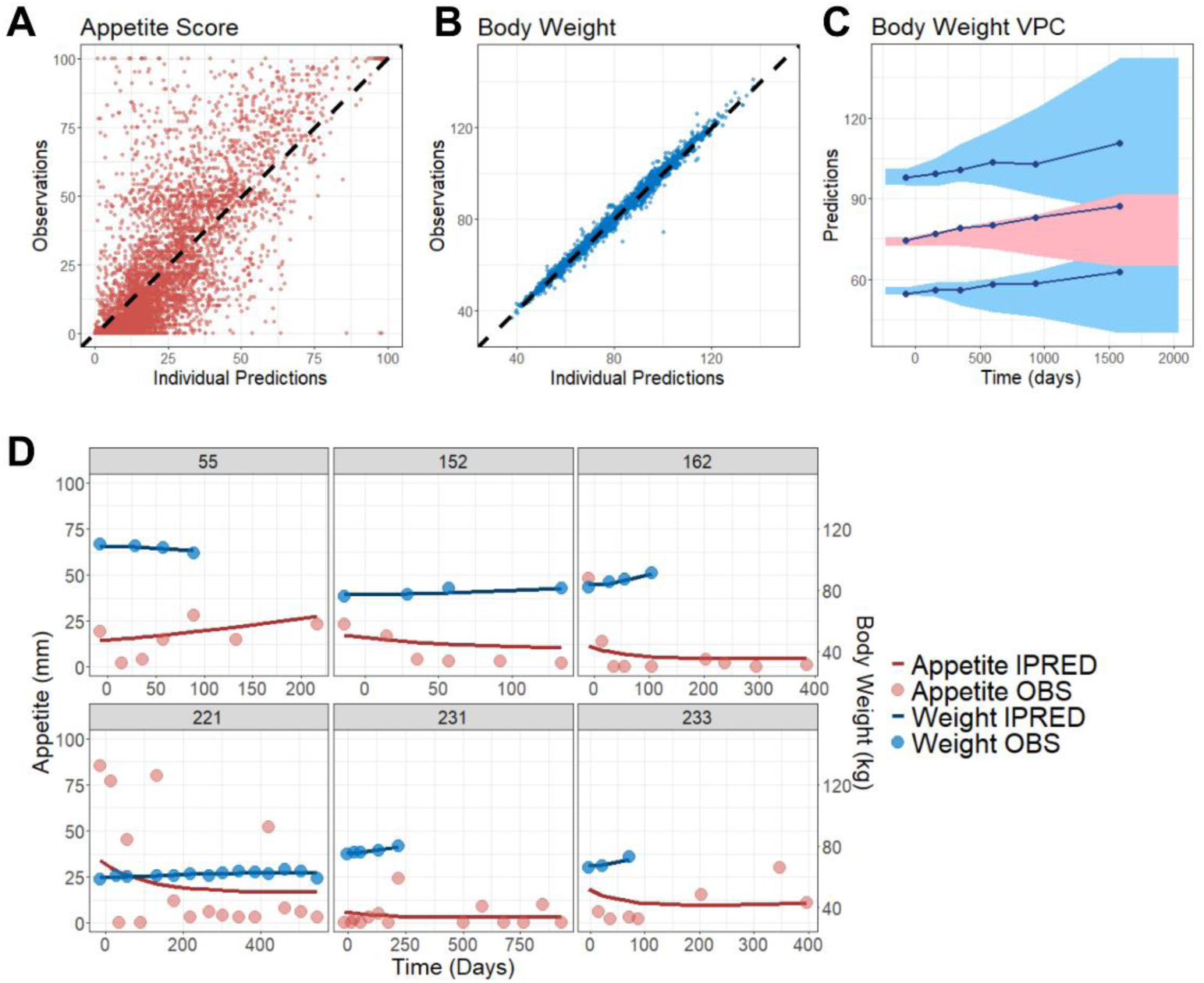
Final model captured appetite and body weight trajectories in recovering study. (A-B) Observations versus individual predictions of appetite scores (A) or body weight (B). The black dashed lines represent the lines of identify. (C) VPC of body weight over time. The observed data are represented by blue solid lines (median and 10th/90th percentiles). The simulated data based on the index population (1000 simulations) are represented by the red shaded area (90% PI of median) or blue shaded area (90% PI of 10th/90th percentiles). VPC has been corrected for dropout. (D) Plots of appetite scores and body weight over time for six randomly selected participants. Circles represent observed data, while solid lines represent individual model predictions. VPC, visual predictive check; PI, prediction interval; IPRED, individual predictions; OBS, observations.

We further validated the model using data from chemotherapy study cohort using same model structure and relationship between appetite and body weight (**Table S1).** The model successfully captured appetite and body weight data in the validation cohort (**Figure S3A-D**), indicating model generalizability across studies. In both the recovering and chemotherapy cohorts, ECOG performance status is a covariate on baseline appetite scores, with higher ECOG scores associated with poorer baseline appetite. Sex was identified as a covariate on baseline body weight, with female patients having 18.8% and 11.3% lower baseline weights than male patients in the recovering and chemotherapy cohorts, respectively. (**Table S1**)

We further compared appetite trajectories between the two cohorts. Patients in the recovering cohort had significantly better baseline appetite (p < 0.0001, **Figure S4A**) and slower appetite decline (p < 0.0001, **Figure S4B**) compared to those in the chemotherapy cohort, which is consistent with expectations that patients tend to lose appetite due to chemotherapy side effects. While the absolute parameter values were beta-transformed and not directly interpretable, the relative differences were consistent with clinical assumptions. [27] Interestingly, the maximum appetite improvement (*PMAX*) was higher in the chemotherapy cohort than in the recovering cohort and showed greater between-subject variability, reflecting the heterogeneous responses and treatment-related toxicities experienced by NSCLC patients during chemotherapy. (**Figure S4C**)

Characterization of the appetite–body weight relationship was achieved by simulating NSCLC population appetite and body weight profiles. (**Figure 3**) During recovery from chemoradiotherapy, NSCLC patient initially experienced appetite improvement, with a 30 mm [90% CI, 28–32] decrease in appetite score (on a 100 mm scale, indicating improved appetite) corresponding to a 3.5 Kg weight gain. Around day 600, the patient’s appetite began to decline, followed by a reduction in body weight, possibly due to disease progression. The time delay between changes in appetite and subsequent changes in body weight forms a hysteresis loop (**Figure 3A**). In contrast, patients under docetaxel chemotherapy experienced persistent appetite loss. A 23 mm [90% CI, 17–30] increase in appetite score was associated with a 3.5 Kg body weight reduction (**Figure 3B**). These findings quantify the appetite–weight relationship in NSCLC and provide a quantitative framework to map appetite loss to corresponding weight loss in future cancer cachexia studies.

**Figure 3.**
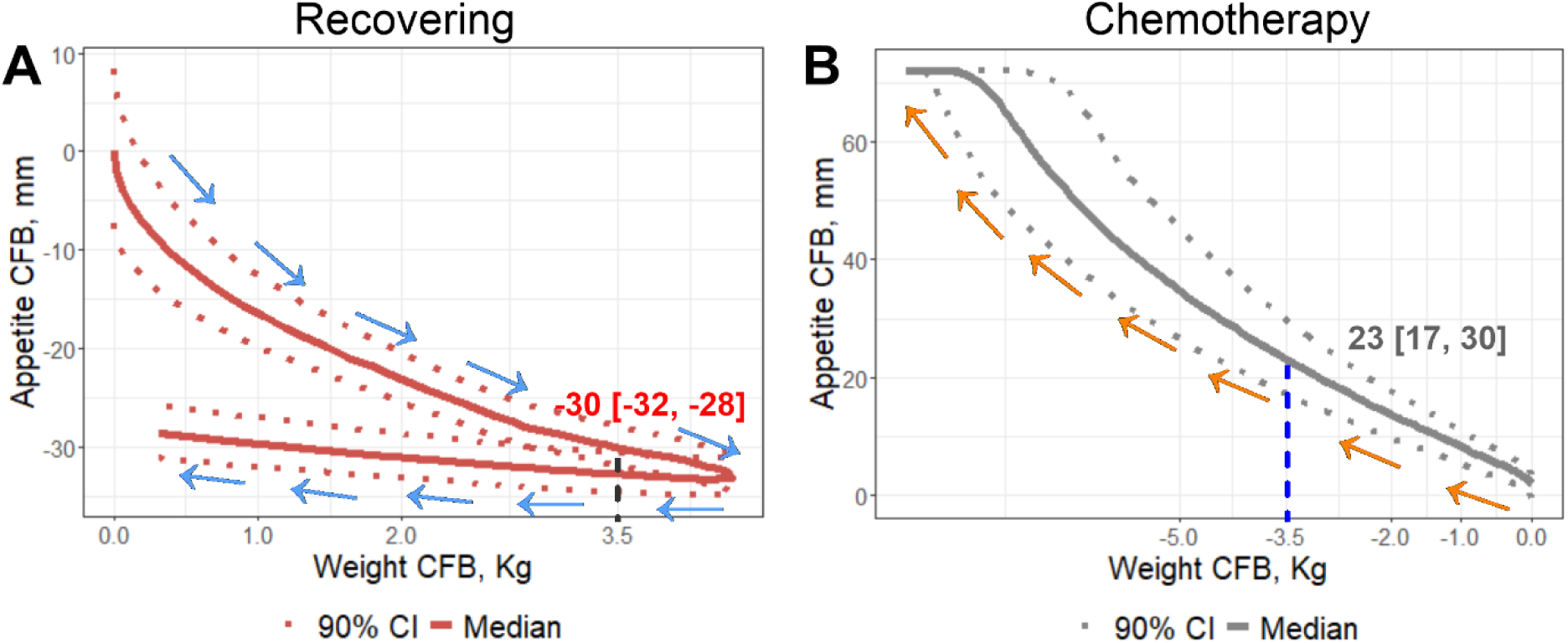
Quantitative relationship between appetite and body weight. (A) In the recovering study, appetite improvement leads to body weight gain, exhibiting a hysteresis loop due to a time delay. A median decrease in appetite score of 30 [90% CI: 28, 32] corresponds to a 3.5 Kg weight increase. (B) In the chemotherapy study, appetite loss leads to body weight reduction. A median increase in appetite score of 23 [90% CI: 17, 30] corresponds to a 3.5 Kg weight loss. CFB, change from baseline; CI, confidence interval; mm, millimeter; Kg, kilogram.

### Linking Appetite Loss Trajectories to OS

To explore the relationship between appetite loss trajectories and OS, Cox proportional hazards models were developed for both recovering and chemotherapy cohorts, incorporating key parameters from the appetite–body weight model. Prognostic covariates were selected using LASSO algorithm. In the recovery cohort, sex was identified as a significant predictor of OS (**Figure S4A**), while in the chemotherapy cohort, smoking status and Eastern Cooperative Oncology Group (ECOG) performance status were identified **(Figure S4B**). After adjusting for these baseline prognostic factors, both *PMAX* (maximal appetite improvements) and *SLP* (appetite loss rate) remained significantly associated with OS. In recovering cohort, a higher appetite loss rate (*SLP*) was associated with poorer survival (HR 1.14, 95% CI: 1.07–1.22), while greater improvements in appetite (*PMAX*) was linked to better survival (HR 0.46, 95% CI: 0.31–0.67). (**Figure 4A**) A similar pattern was observed in the chemotherapy cohort: increased appetite loss was associated with poorer survival (HR 1.22, 95% CI 1.15–1.3) and stronger appetite improvements were associated with improved survival (HR 0.83, 95% CI 0.78–0.88). (**Figure 4B**)

**Figure 4.**
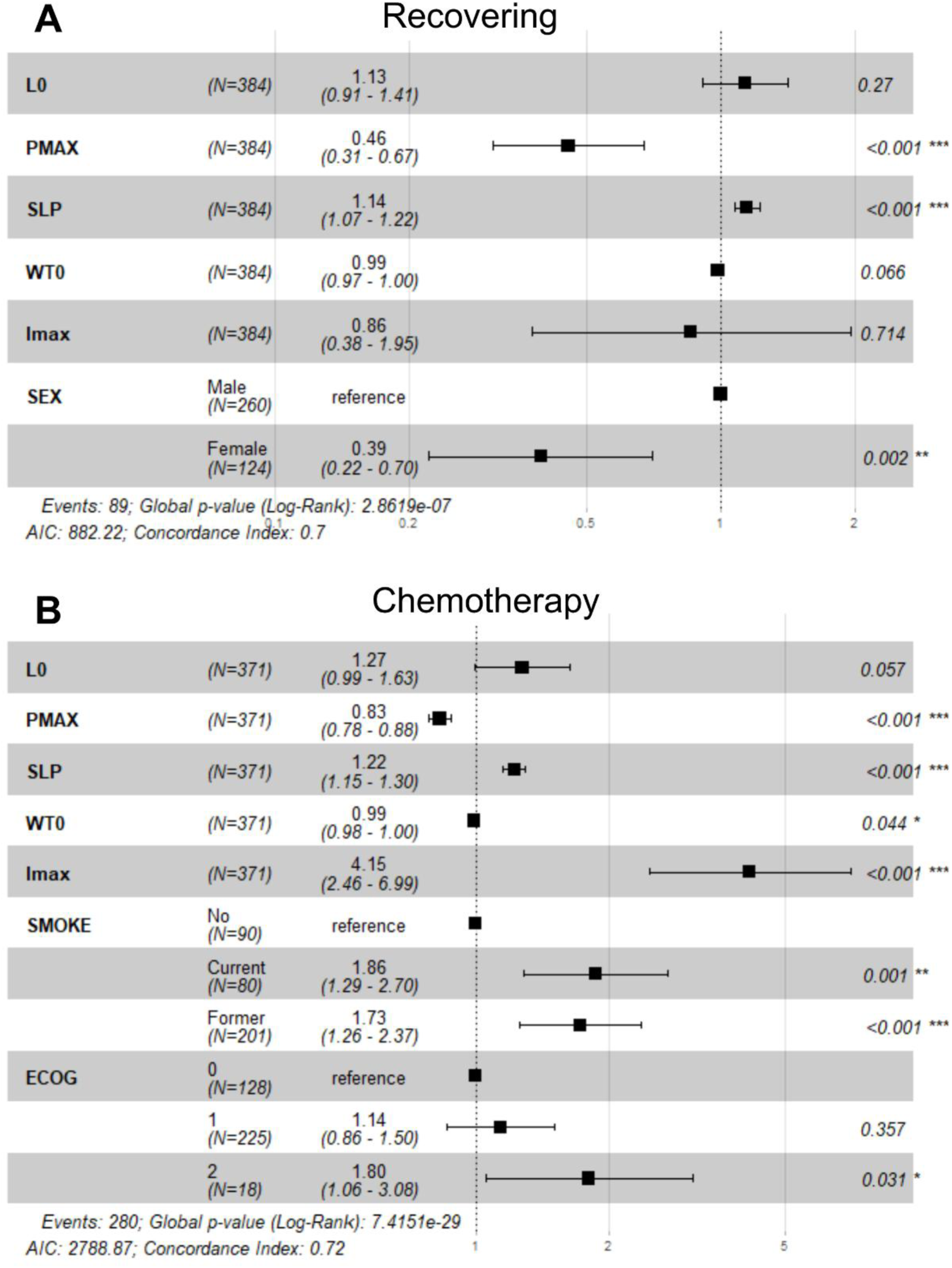
Associations between appetite–body weight model parameters and overall survival. Cox proportional hazards model incorporating model parameters and LASSO-selected survival-related clinical characteristics for recovering study cohort (A) or chemotherapy study cohort (B). The black boxes with horizontal error bars represent hazard ratio estimates with 95% CI. P-values for each covariate are labeled on the right. LASSO, least absolute shrinkage and selection operator; CI, confidence interval; ECOG, Eastern Cooperative Oncology Group. Significance: *, p < 0.05; **, p < 0.01; ***, p < 0.001.

### Identifying Clinically Meaningful Appetite Loss Under Chemotherapy

To identify clinically meaningful appetite loss associated with OS, patients were grouped based on their predicted appetite loss at 1 and 3 months after therapy, with cutoff values ranging from 0 mm to 15 mm tested. Each cutoff, along with prognostic variables identified via LASSO (smoking status and ECOG performance status), was included in Cox proportional hazards models. The resulting hazard ratios and p-values for each cutoff are shown in **Figures 5A** and **5B**. The minimal cutoff associated with p-value < 0.05 in the Cox proportional hazards model— 4 mm at 1 month and 11 mm at 3 months—were selected as clinically meaningful thresholds. Kaplan-Meier curves based on 1 month and 3 months thresholds are shown in **Figures 5C** and **5D**, respectively. Patients with appetite loss ≥ 4 mm at 1 month or ≥ 11 mm at 3 months had significantly worse OS (p < 0.05) compared to those with less appetite loss. Body weight reductions of 1 Kg at 1 month and 3.5 Kg at 3 months were identified as clinically meaningful body weight loss thresholds. Patients who exceeded these thresholds had significantly worse OS compared to those who did not. (p < 0.05, **Figure S5A-D**)

**Figure 5.**
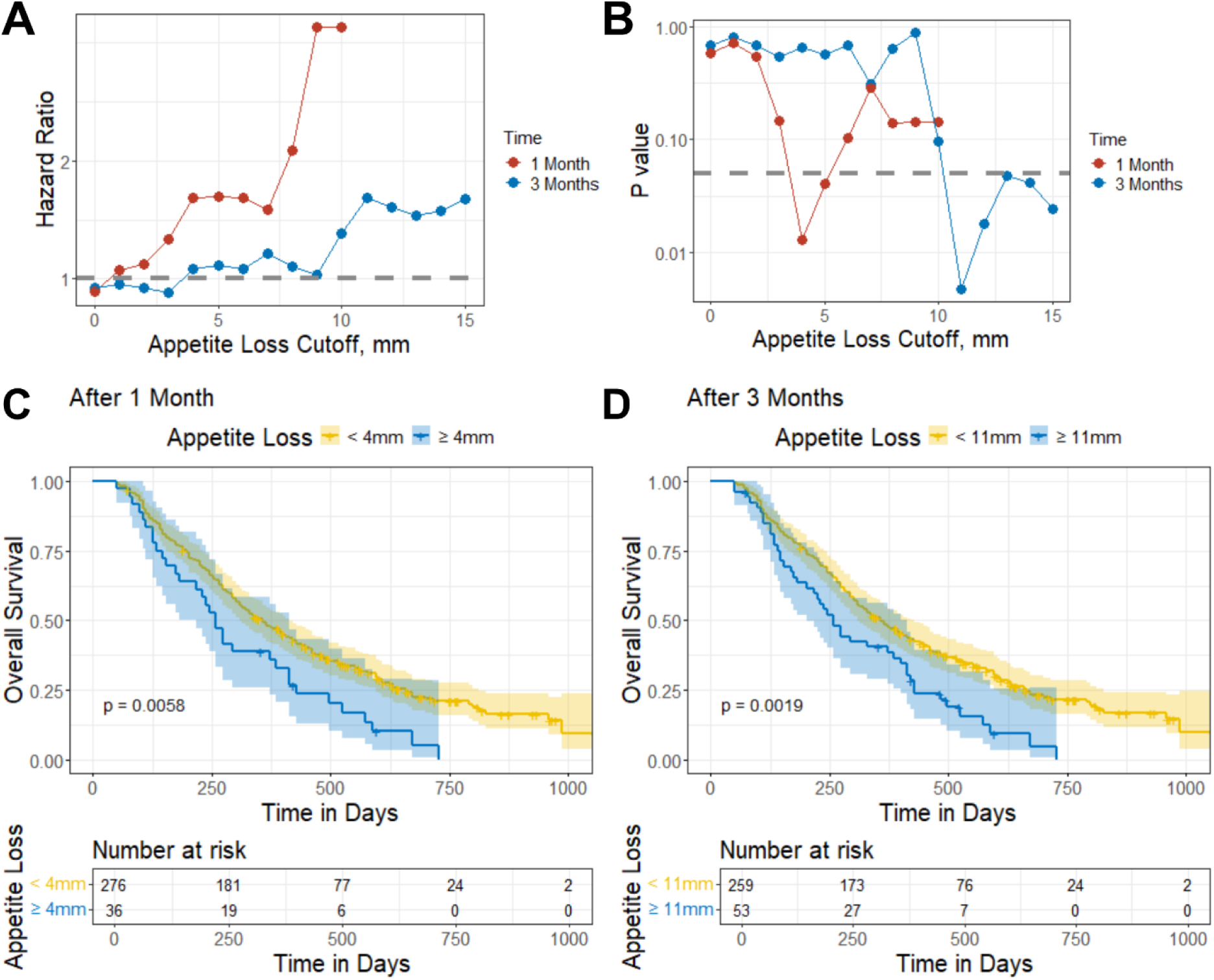
Clinically meaningful thresholds of appetite loss associated with overall survival in chemotherapy study cohort. (A–B) Hazard ratios (A) and p-values (B) from Cox proportional hazards models assessing overall survival across varying appetite loss cutoffs after 1 or 3 months of chemotherapy. (C–D) Kaplan-Meier curves stratified by individual model-predicted appetite loss using a cutoff of 4 mm at 1 month (C) and 11 mm at 3 months (D). Shaded areas indicate 95% confidence intervals; p-values are from log-rank tests. mm, millimeter.

## Discussions

In this study, we developed a novel population modeling approach to quantify the relationship between patient-reported appetite and body weight. On a 100 mm scale, a 30 mm improvement in appetite was associated with a 3.5 Kg weight gain in the recovering cohort, while a 23 mm decline corresponded to a 3.5 Kg weight loss in the chemotherapy cohort. Appetite loss trajectories were also significantly associated with OS: greater loss of appetite was related to worse survival, whereas greater improvement was linked to better outcomes. We identified clinically meaningful appetite loss thresholds—4 mm at 1 month and 11 mm at 3 months—that were significantly associated with OS in NSCLC patients receiving docetaxel chemotherapy. These thresholds may serve as early indicators of cancer cachexia risk and offer a quantitative basis for endpoint selection in future clinical trials.

In our study, we identified clinically meaningful appetite loss based on its significant association with worse overall survival (p < 0.05). The threshold we identified—an 11-point decline after 3 months—is consistent with the commonly used 10-point threshold in the LCSS Average Symptom Burden Index (ASBI) at 12 weeks in lung cancer clinical studies. [28, 29] Notably, our approach utilized a mechanism-based model to define clinically meaningful appetite loss, offering a more sensitive and accurate alternative to traditional empirical endpoint selection. This provides a feasible and rational framework for selecting symptom-based endpoints in future NSCLC clinical trials. In addition, our appetite–body weight model is generalizable across different NSCLC trial populations and can predict individual appetite loss trajectories and the corresponding body weight reduction based on early symptom data. This predictive framework offers the potential to identify patients at higher risk for developing cancer cachexia early in the treatment course.

The appetite–body weight model was initially developed using data from a cohort of patients recovering from chemoradiotherapy and was successfully validated in an independent cohort receiving docetaxel chemotherapy. Notably, patients in the recovery cohort showed better baseline appetite and slower appetite decline, underscoring the positive impact of supportive care in oncology. [30, 31] In contrast, the greater between-subject variability in the maximal appetite improvement parameter observed in the chemotherapy cohort reflected the previous reported heterogeneity in treatment response and toxicity during chemotherapy. [32, 33]

Our study has limitations. First, while appetite loss beyond the identified thresholds is expected to be associated with even smaller p-values given a sufficient sample size, the current analysis shows a "bouncing back" trend in p-values (**Figure 5B**), likely due to the limited sample size in the chemotherapy cohort. Future analyses using larger datasets are needed to validate these thresholds. Second, our findings were derived using the LCSS instrument. For clinical trials that utilize other PRO tools, the differences in scales and response formats would require the development of instrument-specific modeling approaches.

In summary, we developed a population model to characterize the relationship between patient-reported appetite and body weight in NSCLC. We identified the appetite loss thresholds that were associated with 3.5 Kg body weight reduction and significantly worse survival (p < 0.05). These findings provide a quantitative framework to inform symptom-based endpoints in future clinical trials and support early identification of patients at risk for cancer cachexia. By focusing on appetite loss, a frequently overlooked but clinically important symptom, our work supports a patient-centered approach to improve symptom monitoring, guide supportive care strategies, and ultimately enhance the quality of life for patients with NSCLC.

