## Supplementary Materials for "From Symptom to Outcome: Defining Clinically Meaningful Patient-Reported Appetite Loss in Non-Small-Cell Lung Cancer"

### Supplementary Methods

#### 1.1 Population model development

Patient-reported appetite scores and body weight data were modeled simultaneously using nonlinear mixed effect modeling approach with the Stochastic Approximation Expectation-Maximization (SAEM) algorithm. Appetite scores were treated as discrete integer variables bounded between 0 and 100, while body weight was modeled as a continuous variable.

#### 1.2 Bounded data beta-transformation

Many appetite scores in the dataset are reported at the boundaries of the scale (0 or 100). To account for these boundary values, we used an augmented beta distribution (**Equation 1**), which allows for increased probability at the boundaries while modeling the remaining continuous values between 0 and 1 with a beta distribution.

$$p(y_{ij}) = \begin{cases} p0 & if\ y_{ij} = 0 \\ p1 & if\ y_{ij} = 1 \\ (1 - p0 - p1)\frac{\Gamma(\tau)}{\Gamma(\mu_{ij}\tau)\Gamma((1-\mu_{ij})\tau)} {y_{ij}}^{(\mu_{ij}\tau-1)}(1 - y_{ij})^{(1-\mu_{ij})\tau-1} & otherwise \end{cases}$$

(**Equation 1**)

Where $y_{ij}$ is the appetite score reported by subject *i* at time *j*, scaled to a value between 0 and 1. The beta distribution component provides a flexible way to model these values, and is defined using the gamma function ($\Gamma$). The parameter $\mu_{ij}$ represents the expected value of $y_{ij}$, while $\tau$ is the precision parameter that determines how tightly the values cluster around the mean in the gamma function.

To model the probabilities of boundary values ($p0$ and $p1$), we used logistic functions based on the logit transformation of the central value $\mu_{ij}$ (**Equations 2-3**):

$$logit(p0) = -\gamma0 - \gamma1 \times \log\left(\frac{\mu_{ij}}{1-\mu_{ij}}\right)$$ (**Equation 2**)

$$logit(p1) = -\gamma0 + \gamma1 \times \log\left(\frac{\mu_{ij}}{1-\mu_{ij}}\right)$$ (**Equation 3**)

Where $\gamma0$ and $\gamma1$ are the intercept and slope parameters in the beta-transformation that control how the probabilities of observing appetite scores at the boundaries vary with the expected value $\mu_{ij}$.

In summary, this beta-transformation approach provides a flexible framework to model appetite scores across their full range, by combining logistic models for the boundaries with a beta distribution for values between 0 and 1.

**1.3 Random effects model development**

Inter-individual variability (IIV) represents differences in model parameters across individuals. IIV was assumed to follow a normal distribution for the parameters *L0*, *PMAX*, and *SLP* (**Equation 4**), and a log-normal distribution for baseline body weight *WT0* (**Equation 5**). For the *Imax* parameter, a logit-normal distribution was used to constrain values between 0 and 1. No IIV was applied to other parameters, assuming they are shared across all participants in the study.

$$P_i = \theta_P + \eta_i$$ (**Equation 4**)

$$P_i = \theta_P \cdot e^{\eta_i}$$ (**Equation 5**)

Where $P_i$ is the individual value for parameter, $P$, in the $i^{th}$ subject, and $\theta_P$ is the population typical value for parameter $P$, and $\eta$ is an independent random variable describing the variability in $P$ among subjects with a mean of 0 and variance $\omega^2$.

After accounting for IIV, the remaining unexplained variability between observed data and model predictions—potentially due to measurement noise—were captured as residual error. A proportional residual error model was used to describe this variability in observed versus predicted body weight in log scale:

$\log(WT_{ij}) = \log(IPRED_{ij}) \cdot (1 + \varepsilon_{ij,b})$ (**Equation 6**)

Where $WT_{ij}$ is the observed body weight in subject $i$ at time $j$. $IPRED_{ij}$ is the model predicted body weight for subject $i$ at time $j$. $\varepsilon_{ij,b}$ is the proportional error term with mean of 0 and variance of $\sigma^2$.

**1.4 Covariate model development**

Based on exploratory analysis and clinical relevance, potential baseline covariates were evaluated on parameters with IIV (*L0*, *SLP*, *PMAX*, *WT0*, and *Imax*). Covariates for recovering study cohort included age, gender, race, type of initial chemoradiotherapy (concurrent or sequential), previous depression or anxiety history, smoking status, Eastern Cooperative Oncology Group (ECOG) performance status, response to initial chemoradiotherapy (objective response or stable disease). Covariates for chemotherapy study cohort included age, gender, race, previous depression or anxiety history, smoking status, Eastern Cooperative Oncology Group (ECOG) performance status, and disease stage. These covariates were initially plotted against individual parameters to identify any relationships. If a trend was observed, the covariate was included in the full model.

Full fixed effect modeling approach was used for covariate model development. Covariates that show a trend when plotted against individual parameters were added to the base model simultaneously. Correlations between covariates were assessed, and in cases where strong correlations existed, the covariate that was more clinically relevant or with greater statistical significance was selected to be included.

The effect of a categorical covariate on a parameter was represented as a discrete relationship. For example, the effect of ECOG performance status on a parameter *P* was described as exponential effect:

$$P_i = \theta_P \cdot e^{Cov_{ECOG}}, Cov_{ECOG} = \begin{cases} 0 & if\ ECOG\ status = 0 \\ \theta_{ECOG} & if\ ECOG\ status = 1 \end{cases} \text{ (Equation 7)}$$

Where $\theta_{ECOG}$ is the estimable parameter for the effect of ECOG status 1 on parameter *P*.

The effect of a continuous covariate on a parameter was presented as exponential relationship. For example, the effect of Age on a parameter *P* was described as:

$$P_i = \theta_P \cdot e^{\theta_{Age} \cdot Age} \text{ (Equation 8)}$$

Where $\theta_{Age}$ is the estimable parameter for the effect of Age on parameter *P*.

Missing values for covariates were imputed with the population median (for continuous covariates) or mode (for categorical covariates) during the covariate analyses.

### 2. Assessment of Model Performance

#### 2.1 Goodness of Fit

Model goodness-of-fit was assessed by the changes in the minimum objective function value (OFV). Diagnostic plots used to assess model performance included: Observations versus population predictions or individual predictions; Individual weighted residuals versus time or population predictions; Individual predictions over time profiles overlaid with observations.

#### 2.2 Visual predictive check (VPC)

VPC is a population model diagnostic that compares observed data to model-simulated prediction intervals to assess the model's ability to capture variability and central trends in the data. The predictive performance of the final model was evaluated by VPC based on 1000 simulations of the index dataset. In both clinical trials analyzed, we observed substantial participants dropouts from the study due to disease progression or early death. This dropout can cause bias in the VPC and reduce its diagnostic value. To address this, we corrected the VPC for dropout by incorporating dropout predictions into the model and excluding individual predictions occurring after the corresponding predicted dropout time.

#### 2.3 Model Parameters

Two metrics were evaluated to assess model performance: (1) Relative standard error (RSE) reflects the precision of parameter estimates and was reported for all estimated parameters. RSE values below 30% indicate stable and reliable parameter estimation. (2) Shrinkage is the parameter indicating how much individual parameter estimates were pulled towards population typical parameter values and are available for those parameters IIV only. Shrinkage < 40% suggests the model captures sufficient between-subject variability. A high shrinkage (> 60%) means the individual predictions may not be reliable—even when model diagnostics look acceptable overall.

### 3. Model Selection

During the population model development, we evaluated several structural models, including linear and exponential forms of appetite loss, with or without appetite improvement components. Various IIV models were tested, incorporating either log-normal or normal distributions for random effects and applying IIV to different parameters. We also explored multiple residual error models, including additive, proportional, and combined structures. For each candidate model, we assessed diagnostic plots, VPC, and parameter estimates. The final model was selected based on superior performance in goodness-of-fit plots, VPCs, parameter RSE and shrinkage, as well as a lower objective function value.

**4. Model validation**

The model was initially developed using a recovery cohort and externally validated with an independent chemotherapy-treated cohort. The use of a shared model structure and key parameters across both cohorts supports the model's generalizability and robustness.

**Table S1.** Final model parameter estimates.

| | **NCT00409188 (Recovering)** | **NCT00532155 (Chemotherapy)** |
|---|---|---|
| ***Population typical parameter estimates, Value (RSE%)*** | | |
| Baseline appetite score ($\boldsymbol{L0}$)* | -1.46 (5.33%) | -1.14 (7.27%) |
| ECOG performance status 1 covariate effect on baseline appetite ($\boldsymbol{ECOG1\_L0}$)* | 0.335 (30.3%) | 0.267 (32.4%) |
| ECOG performance status 2 covariate effect on baseline appetite ($\boldsymbol{ECOG2\_L0}$)* | Not applicable | 0.436 (47.7%) |
| Maximal appetite improvement ($\boldsymbol{PMAX}$)* | -1.06 (6.99%) | 1.07 (28.3%) |
| Appetite improvement offset rate ($\boldsymbol{Kp}$, 1/day)* | 0.0012 (3.26%)[#] | |
| Appetite loss rate, ($\boldsymbol{SLP}$, 1/day)* | $-3.8\times10^{-3}$ (8.75%) | $2.62\times10^{-3}$ (9.68%) |
| Beta-transformation precision ($\boldsymbol{\tau}$) | 5.67 (2.06%) | 3.65 (5.95%) |
| Beta-transformation slope ($\boldsymbol{\gamma0}$) | 5.82 (2.84%) | 8.09 (9.76%) |
| Beta-transformation intercept ($\boldsymbol{\gamma1}$) | 2.02 (3.47%) | 4.28 (13.3%) |
| Baseline body weight ($\boldsymbol{WT0}$, Kg) | 79.7 (1.25%) | 73.3 (1.25%) |
| Sex (female) covariate effect on baseline body weight ($\boldsymbol{SEX\_WT0}$) | -0.188 (10.4%) | -0.113 (18.2%) |
| Response to initial chemoradiotherapy (stable disease) covariate effect on baseline body weight ($\boldsymbol{BR\_WT0}$) | -0.0649 (30.7%) | Not applicable |
| Zero-order rate constant for appetite loss on body weight reduction ($\boldsymbol{Kin}$, Kg/day) | 1.4 (7%)[#] | |
| Maximal appetite loss impact on body weight reduction ($\boldsymbol{Imax}$, folds) | 0.453 (10.3%)[#] | |
| Appetite loss that reaches 50% Imax body weight reduction ($\boldsymbol{IC50}$) | 12 (3.98%)[#] | |
| ***IIV standard deviation, Value (RSE% \| SHR%)*** | | |
| IIV on baseline appetite score ($\boldsymbol{\Omega_{A0}}$) | 1.04 (3.93% \| 7.5%) | 0.624 (7.84% \| 18.1%) |
| IIV on maximal intervention effect ($\boldsymbol{\Omega_{PMAX}}$) | 0.929 (6.85% \| 46%) | 3.35 (7.18% \| 33.9%) |
| IIV on disease progression rate ($\boldsymbol{\Omega_{SLP}}$) | 4.8 (6.42% \| 21.3%) | 3.09 (10.4% \| 35.7%) |
| IIV on baseline body weight ($\boldsymbol{\Omega_{WT0}}$) | 0.199 (3.3% \| 0.2%) | 0.193 (3.64% \| 0.7%) |
| IIV on maximal appetite loss impact ($\boldsymbol{\Omega_{Imax}}$) | 2.66 (8.5% \| 48.3%) | 2.96 (11.9% \| 57.5%) |

| Residual unexplained variability, Value (RSE%) | | |
|---|---|---|
| Proportional residual error on body weight (***b***) | 0.00537 (1.24%) | 0.00494 (1.46%) |

RSE, relative standard error; IIV, inter-individual variability; SHR, shrinkage.

*Parameter were beta-transformed.

#The RSE% values were obtained from the model for the recovering study. In the chemotherapy study model, parameter values were fixed to those estimated from the recovering model.

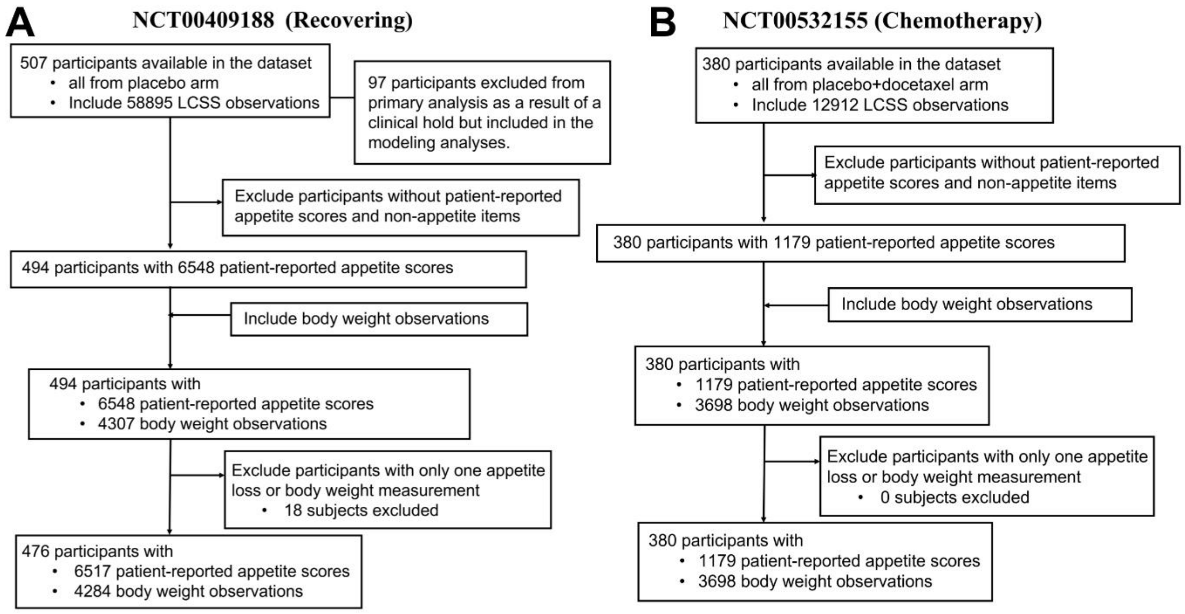


**Figure S1.** Data inclusion and exclusion criteria for recovering dataset NCT00409188 (A) and chemotherapy dataset NCT00532155 (B).

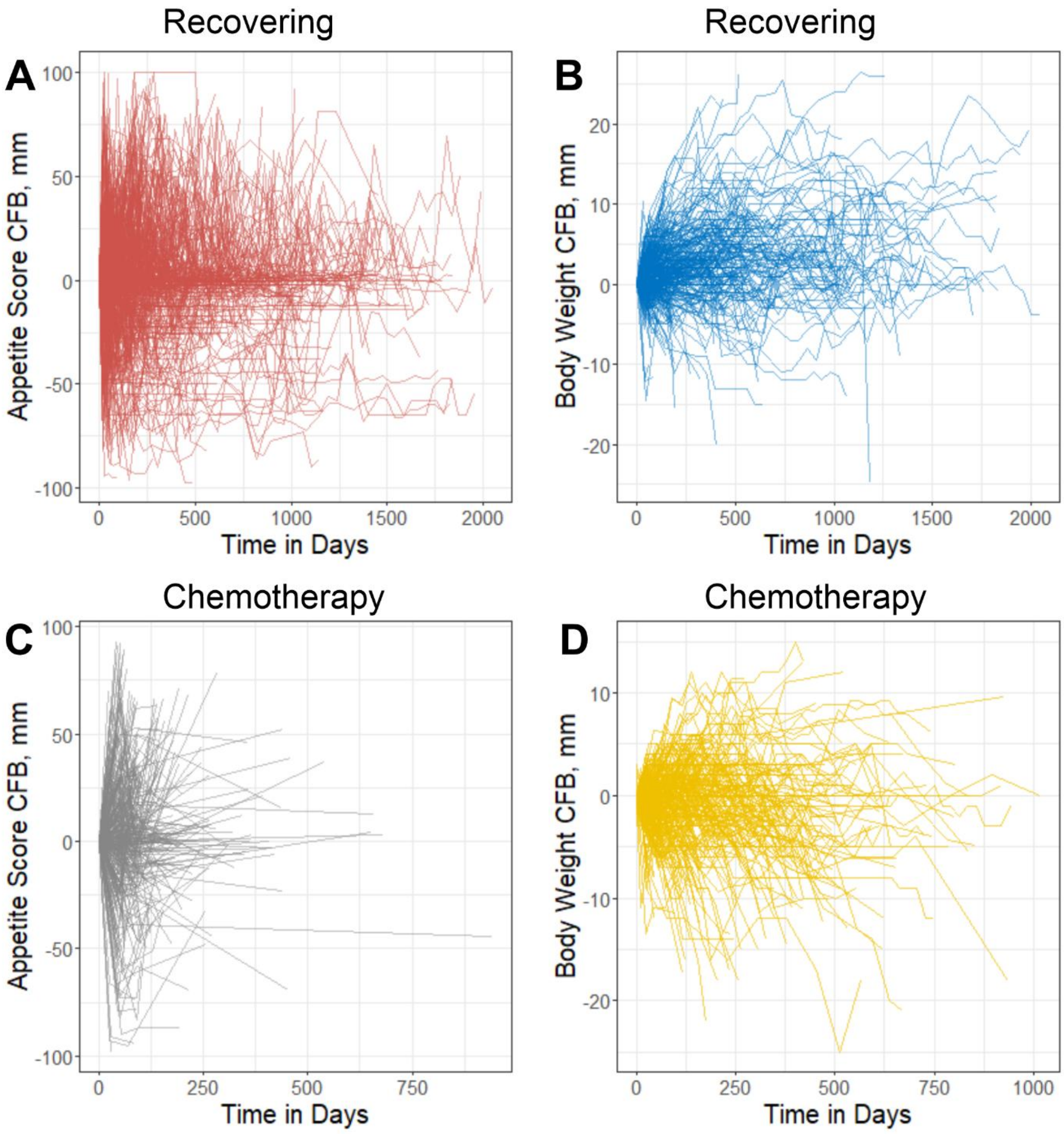


**Figure S2.** Longitudinal appetite score and body weight change from baseline in Recovering study cohort (A-B) and chemotherapy study cohort (C-D). Each line represents one individual patient. CFB, change from baseline.

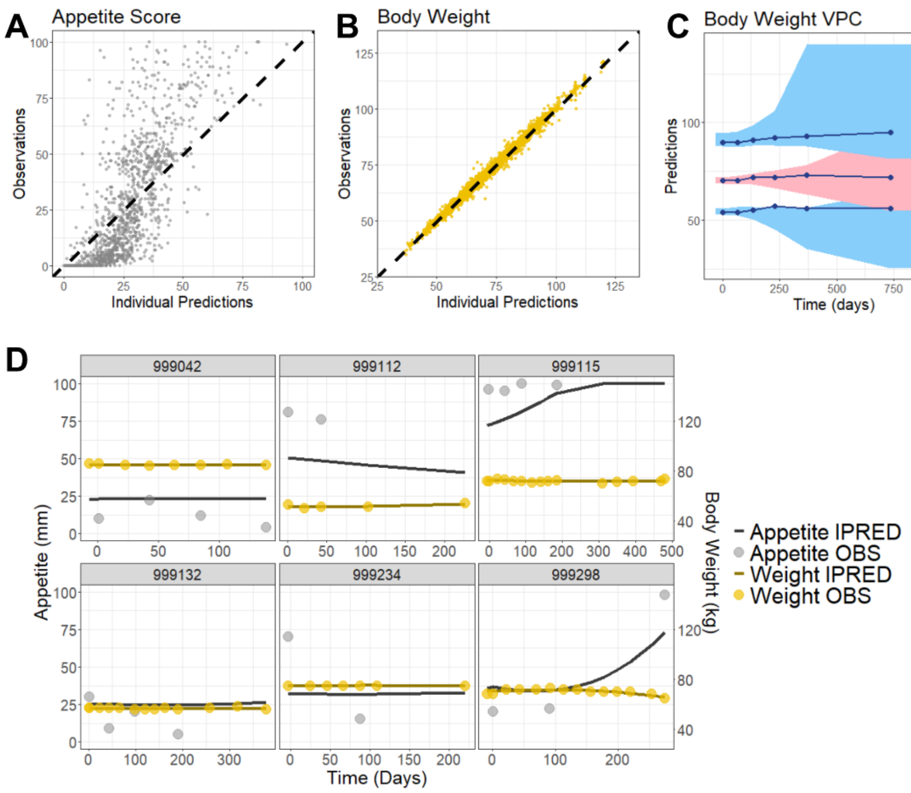


**Figure S3. Final model captured appetite and body weight trajectories in chemotherapy study.** (A-B) Observations versus individual predictions of appetite scores (A) or body weight (B). The black dashed lines represent the lines of identify. (C) VPC of body weight over time. The observed data are represented by blue solid lines (median and 10th/90th percentiles). The simulated data based on the index population (1000 simulations) are represented by the red shaded area (90% PI of median) or blue shaded area (90% PI of 10th/90th percentiles). VPC has been corrected for dropout. (D) Plots of appetite scores and body weight over time for six randomly selected participants. Circles represent observed data, while solid lines represent individual model predictions. VPC, visual predictive check; PI, prediction interval; IPRED, individual predictions; OBS, observations.

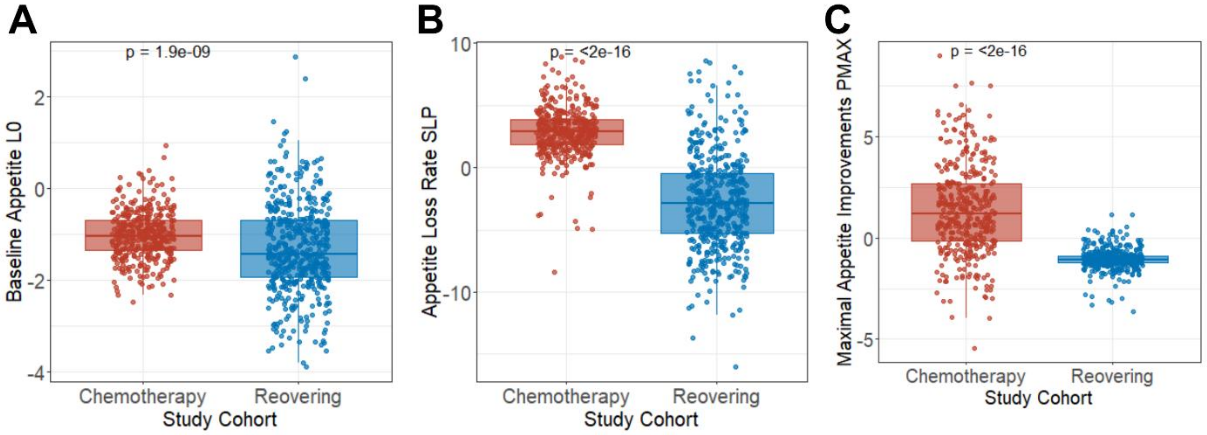


**Figure S4. Boxplots of appetite loss trajectory parameters comparing the recovering and chemotherapy cohorts.** Individual patient-level parameter estimates from the appetite–body weight model are shown for baseline appetite score *L0* (**A**), appetite loss rate *SLP* (**B**), and maximal appetite improvement *PMAX* (**C**). Wilcoxon tests were used to assess differences between cohorts, with p-values indicated above each plot. Boxes represent the interquartile range (IQR), with the horizontal line indicating the median. Whiskers extend to the smallest and largest values within 1.5 times IQR from the lower and upper quartiles; points outside this range are plotted as outliers.

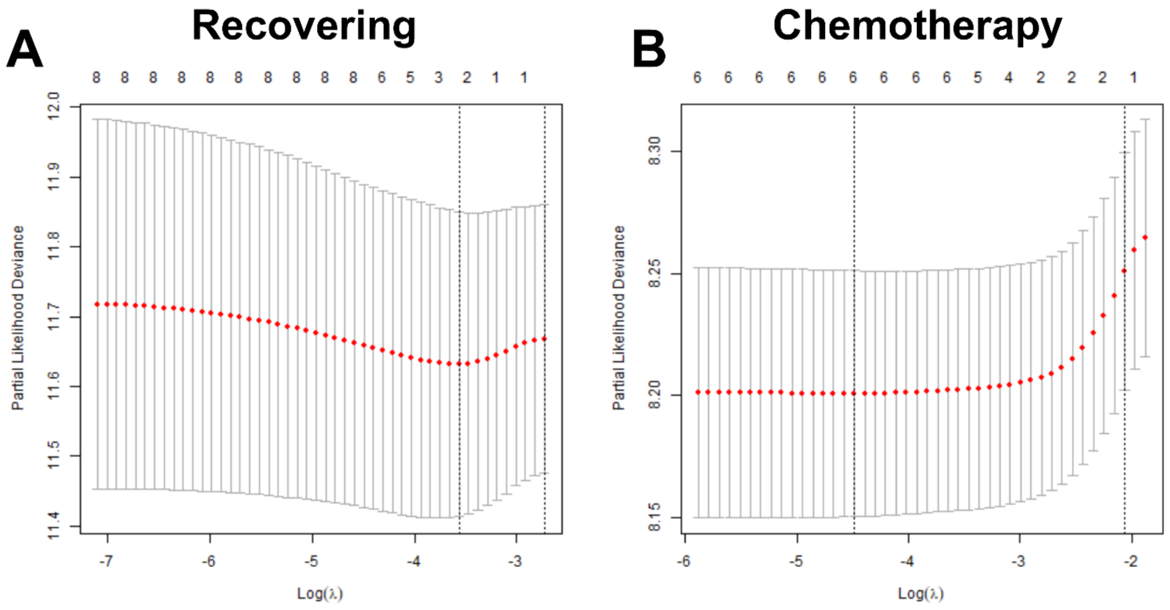


**Figure S5. The relationship between partial likelihood deviance and λ from LASSO algorithms for overall survival in recovering study cohort (A) and chemotherapy study cohort (B).** The red dotted line represents the cross-validation curve, and the gray error bars represent the upper and lower standard deviation curves along the lambda sequence. The selected lambdas (lambda min or lambda se) are indicated by the vertical dashed lines. λ = lamda.se were used to select covariates.

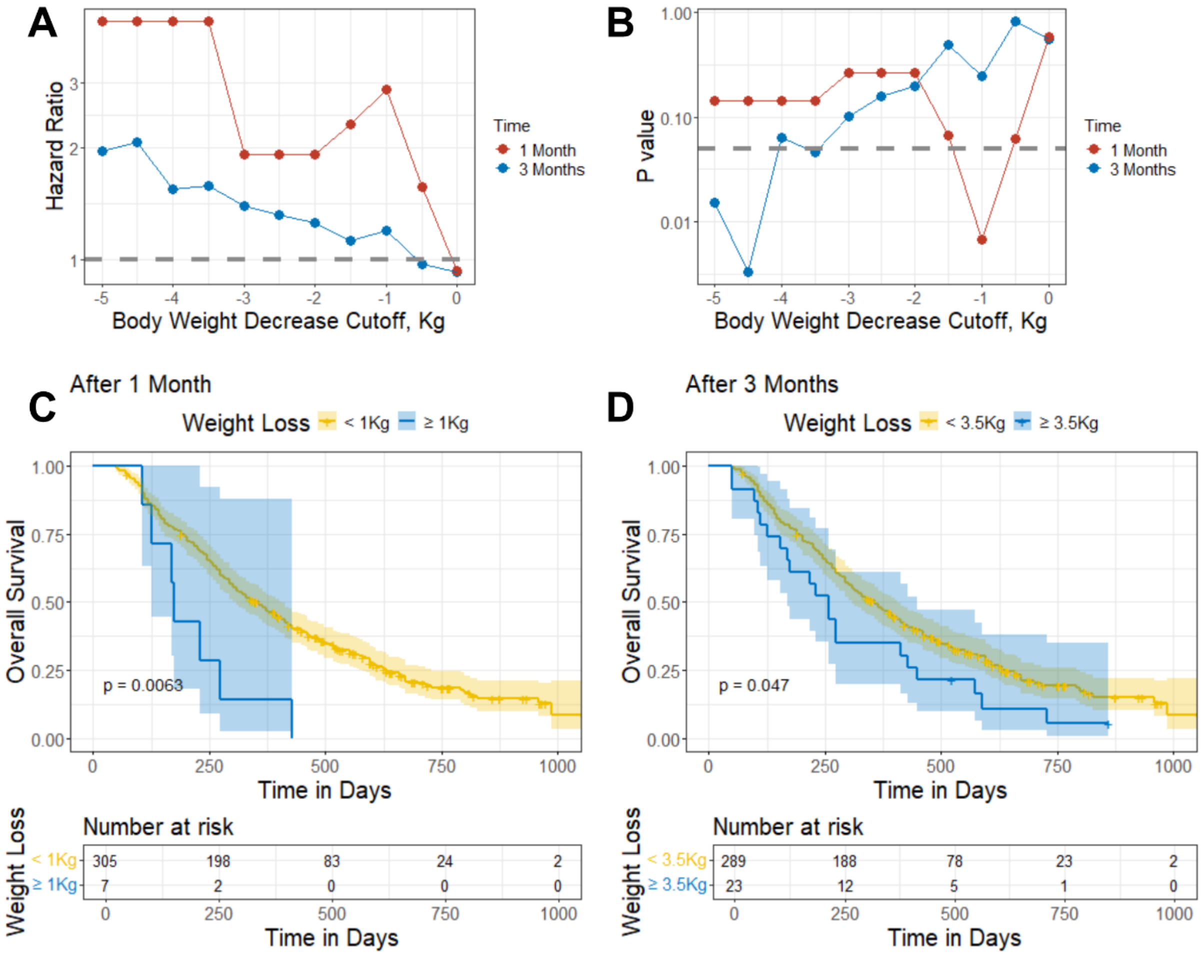


**Figure S6. Clinically meaningful thresholds of body weight loss associated with overall survival in the chemotherapy study.** (A–B) Hazard ratios (A) and p-values (B) from Cox proportional hazards models assessing overall survival across varying body weight loss cutoffs after 1 or 3 months of chemotherapy. (C–D) Kaplan–Meier curves stratified by individual model-predicted body weight loss using a cutoff of 1Kg at 1 month (C) and 3.5Kg at 3 months (D). Shaded areas indicate 95% confidence intervals; p-values are from log-rank tests; Kg, kilogram.

### Supplementary Codes

#### A. Appetite-Body Weight model Monolix Codes

```
DESCRIPTION:
Beta-regression for Appetite score

[LONGITUDINAL]
input={tau, L0, PMAX, Kp, SLP, gamma0, gamma1, FIRSTTIME, WT0, Kin, Imax, IC50}
FIRSTTIME    = {use=regressor}

EQUATION:
; Placebo effects
if t < 0
  PBO = 0
else
  PBO = PMAX * (1 - exp(-Kp * t))
 end

; LCSS mean
SLP2 = SLP * 0.001
DIS = exp(SLP2*(t - FIRSTTIME)) -1
LINP = L0 + DIS - PBO
mu = exp(LINP)/(1+exp(LINP))

maxValue = 100

; LCSS expection
lp0 = -gamma0-gamma1*log(mu/(1-mu))
lp1 = -gamma0+gamma1*log(mu/(1-mu))
p0 = exp(lp0)/(1+exp(lp0))
p1 = exp(lp1)/(1+exp(lp1))
p2 = 1 - p0 - p1
eipred = (p1*1 + p2*mu)*100

; Baseline LCSS
lbmu = L0 + exp(SLP2*(-FIRSTTIME)) - 1
bmu = exp(lbmu)/(1+exp(lbmu))
blp0 = -gamma0-gamma1*log(bmu/(1-bmu))
blp1 = -gamma0+gamma1*log(bmu/(1-bmu))
bp0 = exp(blp0)/(1+exp(blp0))
bp1 = exp(blp1)/(1+exp(blp1))
bp2 = 1 - bp0 - bp1
bipred = (bp1*1 + bp2*bmu)*100

; Body Weight effect
odeType = stiff
t_0 = 0
WT_0 = WT0

TIC50 = IC50*100
bleff = Imax*bipred/(TIC50+bipred)
Kout = Kin*(1-bleff)/WT0
leff = Imax*eipred/(TIC50+eipred)

; weight prediction
ddt_WT = Kin*(1-leff) - Kout*WT


DEFINITION:

LCSSapp = {type=count,
if yij==0
   pk = p0
```

```
elseif yij==maxValue
   pk = p1
else
   pk = (1-p0-p1) * exp(gammaln(tau)-gammaln(mu*tau)-gammaln((1-mu)*tau) + (mu*tau-
1)*log(yij/maxValue) + ((1-mu)*tau-1)*log(1-yij/maxValue) - log(maxValue))
end
P(LCSSapp=yij)= pk }

OUTPUT:
output = {LCSSapp, WT}
table = {p0, p1, p2, mu, eipred, bipred}
```

#### B. Appetite-Body Weight model Monolix Settings for recovering study.

```
<DATAFILE>

[FILEINFO]
file={path='../RecoveringData.csv'}
delimiter = comma
header={C, USUBJID, SUBJID, TIME, DV, QSFLAG, FLAG, MDV, TAFR, AGE, SEX, RACE, ETHNIC, ARM,
PRCANCER, WBC, NEUT, CREAT, BALT, BILI, BALP, BAST, BALB, MHDEPRESS, MHDANXIETY, SMOKE, HEIGHT, BSA,
WEIGHT, ECOG, DTHDY, DTH, PFSDY, PFS, BRESPONSE, FIRSTTIME}

[CONTENT]
USUBJID = {use=identifier}
TIME = {use=time}
DV = {use=observation, yname={'LCSSS1', 'Weight'}, type={discrete, continuous}}
QSFLAG = {use=observationtype}
MDV = {use=missingdependentvariable}
AGE = {use=covariate, type=continuous}
SEX = {use=covariate, type=categorical}
RACE = {use=covariate, type=categorical}
ETHNIC = {use=covariate, type=categorical}
PRCANCER = {use=covariate, type=categorical}
MHDEPRESS = {use=covariate, type=categorical}
MHDANXIETY = {use=covariate, type=categorical}
SMOKE = {use=covariate, type=categorical}
ECOG = {use=covariate, type=categorical}
BRESPONSE = {use=covariate, type=categorical}
FIRSTTIME = {use=regressor}

<MODEL>

[COVARIATE]
input = {AGE, BRESPONSE, ECOG, ETHNIC, MHDANXIETY, MHDEPRESS, PRCANCER, RACE, SEX, SMOKE}

BRESPONSE = {type=categorical, categories={'1', '2'}}
ECOG = {type=categorical, categories={'0', '1'}}
ETHNIC = {type=categorical, categories={'0', '1'}}
MHDANXIETY = {type=categorical, categories={'0', '1'}}
MHDEPRESS = {type=categorical, categories={'0', '1'}}
PRCANCER = {type=categorical, categories={'1', '2'}}
RACE = {type=categorical, categories={'1', '2', '3', '4', '5'}}
SEX = {type=categorical, categories={'0', '1'}}
SMOKE = {type=categorical, categories={'0', '1'}}

[INDIVIDUAL]
input = {Kin_pop, Kp_pop, L0_pop, omega_L0, PMAX_pop, omega_PMAX, SLP_pop, omega_SLP, WT0_pop,
omega_WT0, gamma0_pop, gamma1_pop, tau_pop, IC50_pop, Imax_pop, omega_Imax, ECOG, beta_L0_ECOG_1,
BRESPONSE, beta_WT0_BRESPONSE_2, SEX, beta_WT0_SEX_1}

ECOG = {type=categorical, categories={'0', '1'}}
BRESPONSE = {type=categorical, categories={'1', '2'}}
SEX = {type=categorical, categories={'0', '1'}}

DEFINITION:
Kin = {distribution=logNormal, typical=Kin_pop, no-variability}
Kp = {distribution=logNormal, typical=Kp_pop, no-variability}
L0 = {distribution=normal, typical=L0_pop, covariate=ECOG, coefficient={0, beta_L0_ECOG_1},
sd=omega_L0}
PMAX = {distribution=normal, typical=PMAX_pop, sd=omega_PMAX}
SLP = {distribution=normal, typical=SLP_pop, sd=omega_SLP}
WT0 = {distribution=logNormal, typical=WT0_pop, covariate={BRESPONSE, SEX}, coefficient={{0,
beta_WT0_BRESPONSE_2}, {0, beta_WT0_SEX_1}}, sd=omega_WT0}
gamma0 = {distribution=logNormal, typical=gamma0_pop, no-variability}
gamma1 = {distribution=logNormal, typical=gamma1_pop, no-variability}
```

```
tau = {distribution=logNormal, typical=tau_pop, no-variability}
IC50 = {distribution=logitNormal, typical=IC50_pop, no-variability}
Imax = {distribution=logitNormal, typical=Imax_pop, sd=omega_Imax}

[LONGITUDINAL]
input = {bWeight}

file = 'test28.txt'

DEFINITION:
yWeight = {distribution=logNormal, prediction=WT, errorModel=proportional(bWeight)}

<FIT>
data = {'LCSSS1', 'Weight'}
model = {LCSSapp, yWeight}

<PARAMETER>
IC50_pop = {value=0.1, method=MLE}
Imax_pop = {value=0.2, method=MLE}
Kin_pop = {value=0.2, method=MLE}
Kp_pop = {value=0.0005, method=MLE}
L0_pop = {value=-2, method=MLE}
PMAX_pop = {value=-1, method=MLE}
SLP_pop = {value=-3.5, method=MLE}
WT0_pop = {value=75, method=MLE}
bWeight = {value=0.3, method=MLE}
beta_L0_ECOG_1 = {value=0, method=MLE}
beta_WT0_BRESPONSE_2 = {value=0, method=MLE}
beta_WT0_SEX_1 = {value=0, method=MLE}
cWeight = {value=1, method=FIXED}
gamma0_pop = {value=6, method=MLE}
gamma1_pop = {value=2, method=MLE}
omega_Imax = {value=1, method=MLE}
omega_L0 = {value=1, method=MLE}
omega_PMAX = {value=1, method=MLE}
omega_SLP = {value=1, method=MLE}
omega_WT0 = {value=1, method=MLE}
tau_pop = {value=6, method=MLE}

<MONOLIX>

[TASKS]
populationParameters()
individualParameters(method = {conditionalMean, conditionalMode })
fim(method = StochasticApproximation)
logLikelihood(method = ImportanceSampling)

[PLOTS]
run = true
plots = {indfits = {selected = true}, parameterdistribution = {selected = true}, obspred = {selected
= true}, covariancemodeldiagnosis = {selected = true}, covariatemodeldiagnosis = {selected = true},
vpc = {selected = true}, residualsscatter = {selected = true}, residualsdistribution = {selected =
true}, randomeffects = {selected = true}, saemresults = {selected = true}}

[SETTINGS]
GLOBAL:
exportpath = 'test28'
```

### C. Appetite-Body Weight model Monolix Settings for chemotherapy study.

```
<DATAFILE>

[FILEINFO]
file={path='../ChemotherapyData.csv'}
delimiter = comma
header={RUSUBJID, SUBJID, AGE, SEX, RACE, REGION, ARM, STAGE, DTH, DTHDY, PFS, PFSDY, WBC, NEUT,
CREAT, ALT, BILI, ALP, AST, ALB, MHDEPRESSION, MHANXIETY, SUOCCUR, HEIGHT, WEIGHT, ECOG, TIME, DV,
FLAG, QSTESTCD, QSTEST, B_ASBI, B_LCSS1, B_LCSS2, B_LCSS3, B_LCSS4, B_LCSS5, B_LCSS6, B_LCSS7,
B_LCSS8, B_LCSS9, B_LCSSTOT, TAFR, FIRSTTIME}

[CONTENT]
SUBJID = {use=identifier}
AGE = {use=covariate, type=continuous}
SEX = {use=covariate, type=categorical}
RACE = {use=covariate, type=categorical}
STAGE = {use=covariate, type=categorical}
MHDEPRESSION = {use=covariate, type=categorical}
MHANXIETY = {use=covariate, type=categorical}
SUOCCUR = {use=covariate, type=categorical}
ECOG = {use=covariate, type=categorical}
TIME = {use=time}
DV = {use=observation, yname={'APPETITE', 'WEIGHT'}, type={discrete, continuous}}
QSTEST = {use=observationtype}
FIRSTTIME = {use=regressor}

<MODEL>

[COVARIATE]
input = {AGE, ECOG, MHANXIETY, MHDEPRESSION, RACE, SEX, STAGE, SUOCCUR}

ECOG = {type=categorical, categories={'0', '1', '2'}}
MHANXIETY = {type=categorical, categories={'0', '1'}}
MHDEPRESSION = {type=categorical, categories={'0', '1'}}
RACE = {type=categorical, categories={'1', '5'}}
SEX = {type=categorical, categories={'0', '1'}}
STAGE = {type=categorical, categories={'-999', '1', '2', '3', '4'}}
SUOCCUR = {type=categorical, categories={'0', '1', '2'}}

[INDIVIDUAL]
input = {IC50_pop, Imax_pop, omega_Imax, Kin_pop, Kp_pop, L0_pop, omega_L0, PMAX_pop, omega_PMAX,
SLP_pop, omega_SLP, WT0_pop, omega_WT0, gamma0_pop, gamma1_pop, tau_pop, ECOG, beta_L0_ECOG_1,
beta_L0_ECOG_2, SEX, beta_WT0_SEX_1}

ECOG = {type=categorical, categories={'0', '1', '2'}}
SEX = {type=categorical, categories={'0', '1'}}

DEFINITION:
IC50 = {distribution=logitNormal, typical=IC50_pop, no-variability}
Imax = {distribution=logitNormal, typical=Imax_pop, sd=omega_Imax}
Kin = {distribution=logNormal, typical=Kin_pop, no-variability}
Kp = {distribution=logNormal, typical=Kp_pop, no-variability}
L0 = {distribution=normal, typical=L0_pop, covariate=ECOG, coefficient={0, beta_L0_ECOG_1,
beta_L0_ECOG_2}, sd=omega_L0}
PMAX = {distribution=normal, typical=PMAX_pop, sd=omega_PMAX}
SLP = {distribution=normal, typical=SLP_pop, sd=omega_SLP}
WT0 = {distribution=logNormal, typical=WT0_pop, covariate=SEX, coefficient={0, beta_WT0_SEX_1},
sd=omega_WT0}
gamma0 = {distribution=logNormal, typical=gamma0_pop, no-variability}
gamma1 = {distribution=logNormal, typical=gamma1_pop, no-variability}
tau = {distribution=logNormal, typical=tau_pop, no-variability}

[LONGITUDINAL]
```

```
input = {bWEIGHT}

file = 'test3.txt'

DEFINITION:
yWEIGHT = {distribution=logNormal, prediction=WT, errorModel=proportional(bWEIGHT)}

<FIT>
data = {'APPETITE', 'WEIGHT'}
model = {LCSSapp, yWEIGHT}

<PARAMETER>
IC50_pop = {value=0.12, method=FIXED}
Imax_pop = {value=0.453, method=FIXED}
Kin_pop = {value=1.4, method=FIXED}
Kp_pop = {value=0.0012, method=FIXED}
L0_pop = {value=-2, method=MLE}
PMAX_pop = {value=-1, method=MLE}
SLP_pop = {value=1, method=MLE}
WT0_pop = {value=70, method=MLE}
bWEIGHT = {value=0.3, method=MLE}
beta_L0_ECOG_1 = {value=0, method=MLE}
beta_L0_ECOG_2 = {value=0, method=MLE}
beta_WT0_SEX_1 = {value=0, method=MLE}
cWEIGHT = {value=1, method=FIXED}
gamma0_pop = {value=6, method=MLE}
gamma1_pop = {value=2, method=MLE}
omega_Imax = {value=1, method=MLE}
omega_L0 = {value=1, method=MLE}
omega_PMAX = {value=1, method=MLE}
omega_SLP = {value=1, method=MLE}
omega_WT0 = {value=1, method=MLE}
tau_pop = {value=6, method=MLE}

<MONOLIX>

[TASKS]
populationParameters()
individualParameters(method = {conditionalMean, conditionalMode })
fim(method = StochasticApproximation)
logLikelihood(method = ImportanceSampling)

[PLOTS]
run = true
plots = {indfits = {selected = true}, parameterdistribution = {selected = true}, obspred = {selected
= true}, covariancemodeldiagnosis = {selected = true}, covariatemodeldiagnosis = {selected = true},
vpc = {selected = true}, residualsscatter = {selected = true}, residualsdistribution = {selected =
true}, randomeffects = {selected = true}, saemresults = {selected = true}}

[SETTINGS]
GLOBAL:
exportpath = 'test3'
```
